# Impact of chronic pain on numerous long COVID-like symptoms: A large-scale internet-based epidemiological study in Japan

**DOI:** 10.1101/2024.11.11.24317094

**Authors:** Saki Takaoka, Hanako Saito, Morihiko Kawate, Chisato Tanaka, Yihuan Wu, Shizuko Kosugi, Takashige Yamada, Takahiro Tabuchi, Kenta Wakaizumi

## Abstract

**Background:** Individuals with chronic pain (CP) not only endure the direct burden of the pain but also experience various symptoms, including sleep disturbances and fatigue, which deteriorate their quality of life. Notably, these symptoms closely resemble those observed in ‘long COVID’, a prolonged health complication that can arise after coronavirus disease 2019 (COVID-19) infection. However, the similarities between them have not yet been explored. Therefore, this study aimed to investigate the relationship between CP and long COVID using Japanese epidemiological data.

**Methods:** Utilizing the Japan COVID-19 and Society Internet Survey in 2022, an Internet-based survey involving 32,000 participants, we analysed data on the presence of CP, history of COVID-19 infection, and presence of 17 long COVID-like symptoms, including gastrointestinal upset, back pain, limb/joint pain, headache, chest pain, shortness of breath, dizziness, sleep disorder, hearing disorder, taste disorder, smell disorder, memory impairment, poor concentration, hair loss, decreased libido, fatigue, and cough.

**Results:** Individuals with history of COVID-19 experienced a significantly greater number of long COVID-like symptoms (median: 5) compared to those with neither COVID-19 nor CP (median: 4, *p* < 0.001). However, individuals with CP alone and those with both COVID-19 and CP exhibited an even greater number of symptoms (median: 8 and 9, respectively). Additionally, individuals with CP exhibited greater prevalence odds for 15 of the 17 symptoms than those without CP or COVID-19 (*p* < 0.001).

**Conclusions:** Altogether, our study suggests that the long COVID-like symptoms are not specifically related to COVID-19, but CP is an independent risk factor for these symptoms.

## Introduction

Chronic pain (CP) is one of the most common health problems in modern society, affecting approximately 20–40% of the global population.^1^ According to a national survey in Japan, back pain, shoulder pain, joint pain, and headache consistently rank among the top five reported physical concerns for both men and women.^2^

Patients with CP not only endure a substantial burden from the pain but also experience psychological and social impairments associated with the condition, including anxiety, depression, pain-related fear, reduced daily activity, decreased work performance, and social withdrawal.^1,3,4^ Furthermore, individuals with CP experience comorbid physical conditions, including sleep disturbances and fatigue, that further compromise their quality of life.^5–7^ These psychological and physical symptoms of CP share similarities with the complaints reported by patients with ‘long COVID’.

Long COVID is a relatively new disease entity that has emerged as a consequence of the coronavirus disease 2019 (COVID-19). COVID-19 has caused a global pandemic, infecting over 776 million people worldwide.^8^ It is estimated that 10-35 %^9^ of those infected may develop long-term health complications following the acute phase of infection, a condition commonly known as ‘long COVID’ or post COVID-19 condition. Common symptoms of long COVID include fatigue, sleep disturbances, joint pain, anxiety, depression, and cognitive impairment.^10^ However, the relationship between CP and long COVID have not yet been explored. Therefore, this study aimed to investigate their relationship using large-scale epidemiological data from Japan.

## Methods

### Study participants

Data from the Japan COVID-19 and Society Internet Survey (JACSIS) was used for this study. The JACSIS is a large-scale, Internet-based longitudinal survey conducted annually in Japan since 2020. The project is aimed to collect data on the lifestyle, health condition, and socioeconomic activities of Japanese people after the COVID-19 outbreak and use the data to provide health-promoting strategies and socioeconomic relief measures. The survey is supported by a Japanese Internet research agency (Rakuten Insight, Inc., Tokyo, Japan), encompassing 2.2 million nationwide panellists. The panellists received a financial incentive of JPY 100 (approximately USD 0.7 in September 2022) for participating in an Internet-based, self-reported questionnaire survey. For this study, we selected data from the JACSIS 2022 survey, which was conducted between 15 September and 15 October 2022.

### Data collection

Participants were asked to respond to questions concerning their lifestyle, health, and socioeconomic status in the JACSIS 2022 questionnaire. For the present study, we extracted data on the presence of CP, history of COVID-19, presence of 17 long COVID-like symptoms during the previous week, psychological distress levels, comorbidities, and demographic and lifestyle measures.

### Chronic pain

CP was defined as the presence of any pain lasting for more than 3 months. The current or past history of CP among the participants was recorded. Individuals who reported having CP at the time of assessment were classified as having CP.

### History of COVID-19 infection

Participants were asked whether they had been diagnosed with COVID-19 within the past 2 months, between 1 year and 2 months ago, or more than a year ago. Those meeting any of the above conditions were classified as having a history of COVID-19 infection.

### Long COVID-like symptoms

For long COVID-like symptoms, 17 common symptoms, which are gastrointestinal upset, back pain, limb/joint pain, headache, chest pain, shortness of breath, dizziness, sleep disorders, hearing disorders, taste disorders, smell disorders, memory impairment, poor concentration, hair loss, decreased libido, fatigue, and cough, were selected for analysis based on previous reports.^11^ Participants were asked to rate the extent of their distress regarding each symptom experienced in the past week using a 5-point Likert scale, with responses ranging from 1 (not at all) to 5 (very much). A score of 2 (slightly) or higher was considered as the presence of any symptoms.

### Psychological distress levels

The psychological distress levels of the participants were assessed using the Japanese version of the Kessler Psychological Distress Scale (K6), which is a well-validated questionnaire (Cronbach’s = α 0.85).^12–14^ In the JACSIS 2022 questionnaire, participants were asked to rate their psychological distress during the past 30 days for six items of the K6 (e.g., ‘During the past 30 days, how often did you feel nervous?’) on a 5-point scale ranging from 0 (never) to 4 (always). The total score for psychological distress was calculated by summing the scores of the six items, with a higher total score indicating greater psychological distress. Participants with a score of 13 points or higher were classified as having serious psychological distress.^14,15^

### Comorbidities

The participants were asked about the presence of 19 disorders: hypertension, diabetes, hyperlipidaemia, pneumonia/bronchitis, asthma, atopic dermatitis, allergic rhinitis, periodontal disease, dental caries, cataract, angina pectoris/myocardial infarction, stroke, chronic obstructive pulmonary disease, chronic kidney disease, chronic hepatitis/liver cirrhosis, immune disorder, cancer/malignant tumour, depression, and other mental disorders. As multiple comorbidities have been reported as risk factors for long COVID,^16,17^ the total number of comorbidities for each participant was calculated for analysis.

### Demographic and lifestyle-related measures

For demographic and lifestyle-related measures, information was collected on age, sex, body mass index (BMI; kg/m^2^), employment status (executive, employer, self-employed, employee, homemaker, unemployed, retired, or student), marital status (married, never married, widowed, or divorced), highest educational level (high school or lower, or college or higher), annual income in Japanese yen (below the average income in Japan, 4 million yen,^18^ or 4 million yen and above), number of COVID-19 vaccinations, and smoking habits (never, ex-smoker, or current smoker).

### Exclusion criteria

We excluded respondents who met at least one of the following conditions due to inconsistent responses^11^: (1) failed to provide a correct response to an attention check question instructing them to select the penultimate option from the five given options, (2) reported having 15 or more family members, (3) reported the presence of all nine health conditions listed, including hypertension, diabetes, asthma, atopic dermatitis, allergic rhinitis, cataract, angina pectoris/myocardial infarction, immune disorder, and cancer/malignant tumour, and (4) claimed to have used all five categories of illegal drugs listed, including narcotics (e.g., morphine) that are not obtained through a doctor’s prescription, organic solvents (e.g., thinner and toluene), designer drugs (e.g., synthetic herbs and magic mushrooms), cannabis, and stimulants, cocaine, and heroin.

### Statistical analysis

The JACSIS 2022 survey participants were classified into the following four groups based on the presence of CP and history of COVID-19: (1) without CP or COVID-19, (2) with COVID-19 alone, (3) with CP alone, and (4) with both CP and COVID-19. The total number of reported long COVID-like symptoms was compared among the four groups using the Kruskal–Wallis test, followed by the Steel–Dwass test for post hoc analysis, as a nonparametric multiple comparison test. To eliminate the influence of pain-related symptoms, we repeated the analysis after excluding four pain-related symptoms: back pain, limb/joint pain, headache, and chest pain. The prevalence of each long COVID-like symptom was compared among the four groups using the chi-square test. The Bonferroni correction was applied for post hoc comparisons. Odds ratios for the prevalence of each long COVID-like symptom were computed to assess the effects of COVID-19 alone, CP alone, and both COVID-19 and CP using multivariable logistic regression analysis. Potential confounding factors associated with chronic pain, long COVID, or both, including age, sex, BMI, employment status, marital status, highest educational level, income, number of comorbidities, psychological distress, number of COVID-19 vaccinations, and smoking habits, were adjusted for the analysis. All statistical analyses were performed using the JMP^®^ Pro Version 17.2.0 (SAS Institute Inc., Cary, NC, USA). Statistical significance was set at two-tailed *p* < 0.05.

### Ethical approval

All procedures were approved by the Keio University School of Medicine Ethics Committee (approval number: 20230001) and the Ethics Review Committee of the Osaka International Cancer Centre, Osaka Prefectural Hospital Organization (approval number: 20084).

### Informed consent

All participants were informed about the aim of the survey through the Rakuten Insight webpage. All participants provided informed consent prior to completing the questionnaire.

## Results

Among the 32,000 panellists who participated in the JACSIS 2022 survey, 3,389 (10.6%) with inconsistent responses were excluded. Subsequently, 28,611 respondents were included in the analysis.

Table 1 summarises the demographic characteristics of the participants in the four groups. In total, 20,184 participants (70.5%) reported having neither of the two conditions, 4,286 (15.0%) reported having CP alone, 3,360 (11.7%) reported having a history of COVID-19, and 781 (2.7%) reported having both CP and a history of COVID-19. The total number of people who reported to have CP was 5,067 (17.7 %). Individuals with CP alone were older (mean age: 52.8 years) and had lower incomes compared with the other three groups. Furthermore, the proportion of males (44.2%) was lower among the individuals with CP alone. Individuals with both CP and COVID-19 had more comorbidities and a greater prevalence of psychological distress (24.7%) than the other three groups.

**Table 1.**
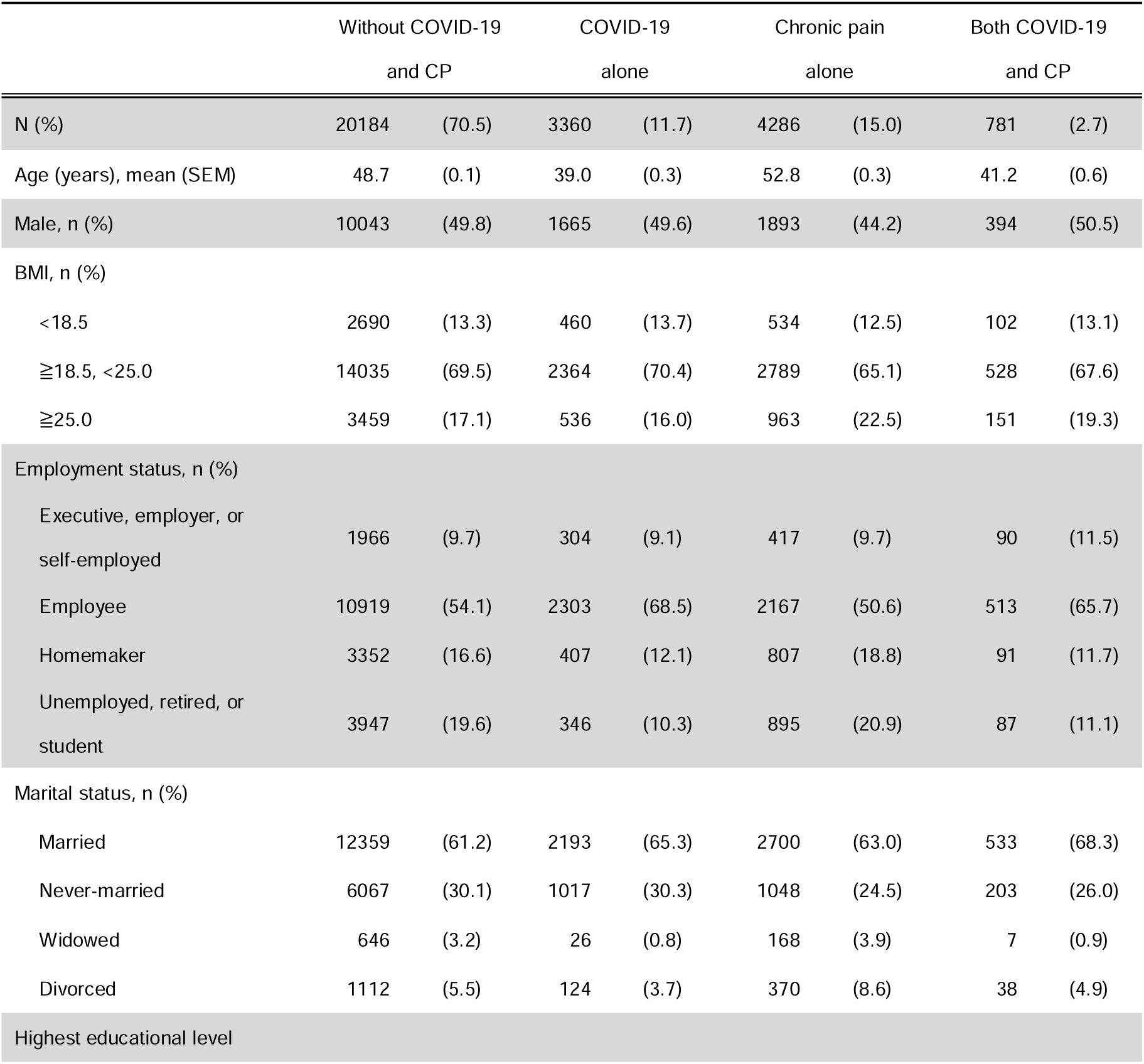
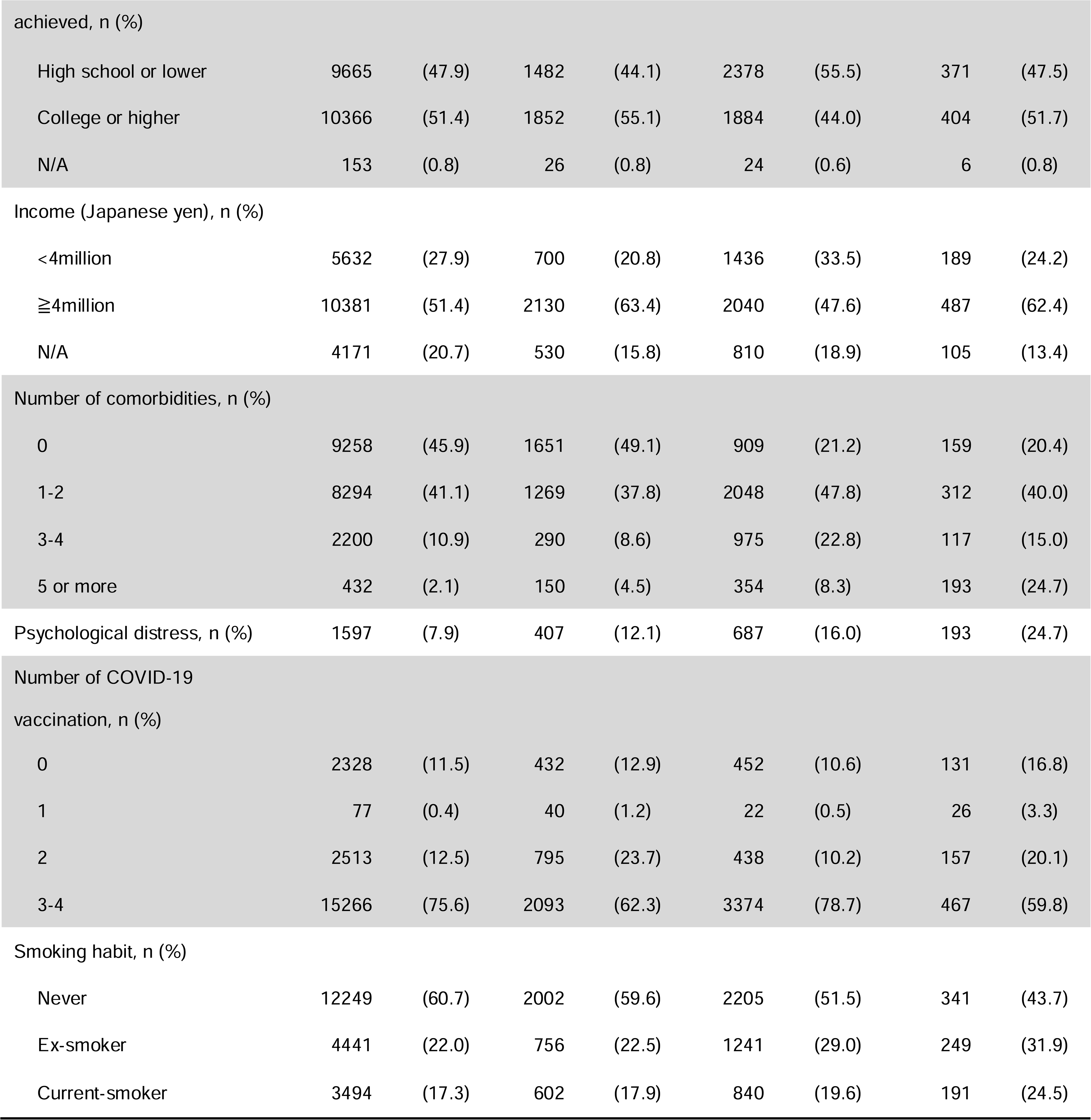
Demographic characteristics of participants across the four groups based on the presence of chronic pain and the history of COVID-19 infection (N = 28,611). BMI: body mass index, CKD: chronic kidney disease, COPD: chronic obstructive pulmonary disease, SEM: standard error of the mean.

Figure 1A presents the total number of long COVID-like symptoms reported for each group. A significant increase was observed in the number of symptoms across the groups in the following order: without COVID-19 and CP (median number of symptoms: 4), COVID-19 alone (median: 5), CP alone (median: 8), and both COVID-19 and CP (median: 9). Furthermore, a significantly greater number of symptoms was observed in patients with COVID-19 alone, CP alone, and both COVID-19 and CP, even when pain-related symptoms were excluded (Figure 1B).

**Figure 1.**
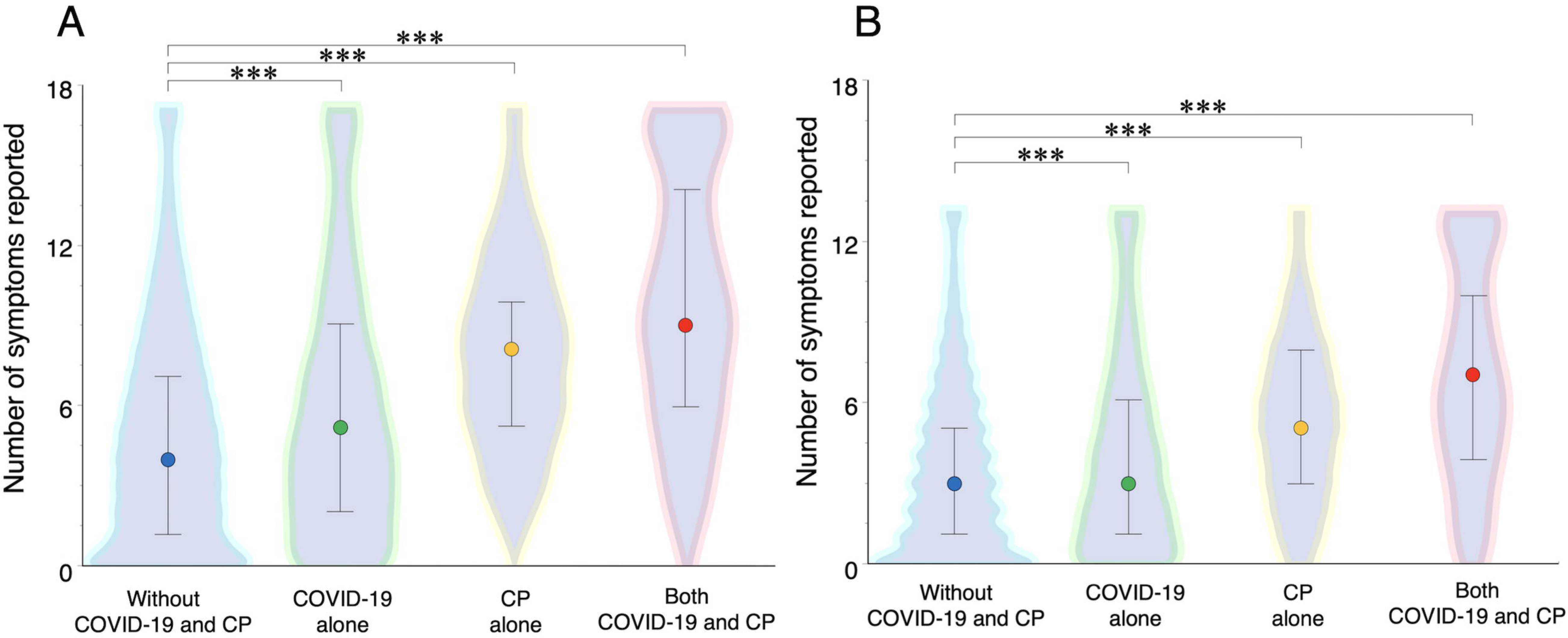
Comparison of the number of reported long COVID-like symptoms (N = 28,611) across the groups. A) The total number of the symptoms was compared among the four groups using the Kruskal–Wallis test, followed by Steel–Dwass post hoc analysis. B) The total number of symptoms excluding the four pain-related symptoms (back pain, limb/joint pain, headache, and chest pain) was compared among the four groups. Circle represents the median value, and bar represents the interquartile range. \*\*\**p* < 0.001.

Figure 2 presents the prevalence of each long COVID-like symptom across the four groups. The prevalence increased in the order of groups without COVID-19 and CP, with COVID-19 alone, with CP alone, and with both COVID-19 and CP for the following 12 symptoms: gastrointestinal upset, headache, chest pain, shortness of breath, dizziness, sleep disorder, hearing disorder, memory impairment, poor concentration, hair loss, decreased libido, and fatigue. In contrast, individuals with CP alone exhibited the highest prevalence of two pain-related symptoms, which are back pain and limb/joint pain. Furthermore, individuals with CP alone exhibited a higher prevalence of certain respiratory symptoms, including chest pain and shortness of breath, and certain central nervous system symptoms, including dizziness, sleep disorder, memory impairment, poor concentration, and fatigue, than those with COVID-19 alone. On the other hand, individuals with COVID-19 alone exhibited a higher prevalence of taste disorder, smell disorder, and cough than those with CP alone. Individuals with both CP and COVID-19 exhibited the highest prevalence of all symptoms, except for back pain and limb/joint pain, among the four groups.

**Figure 2.**
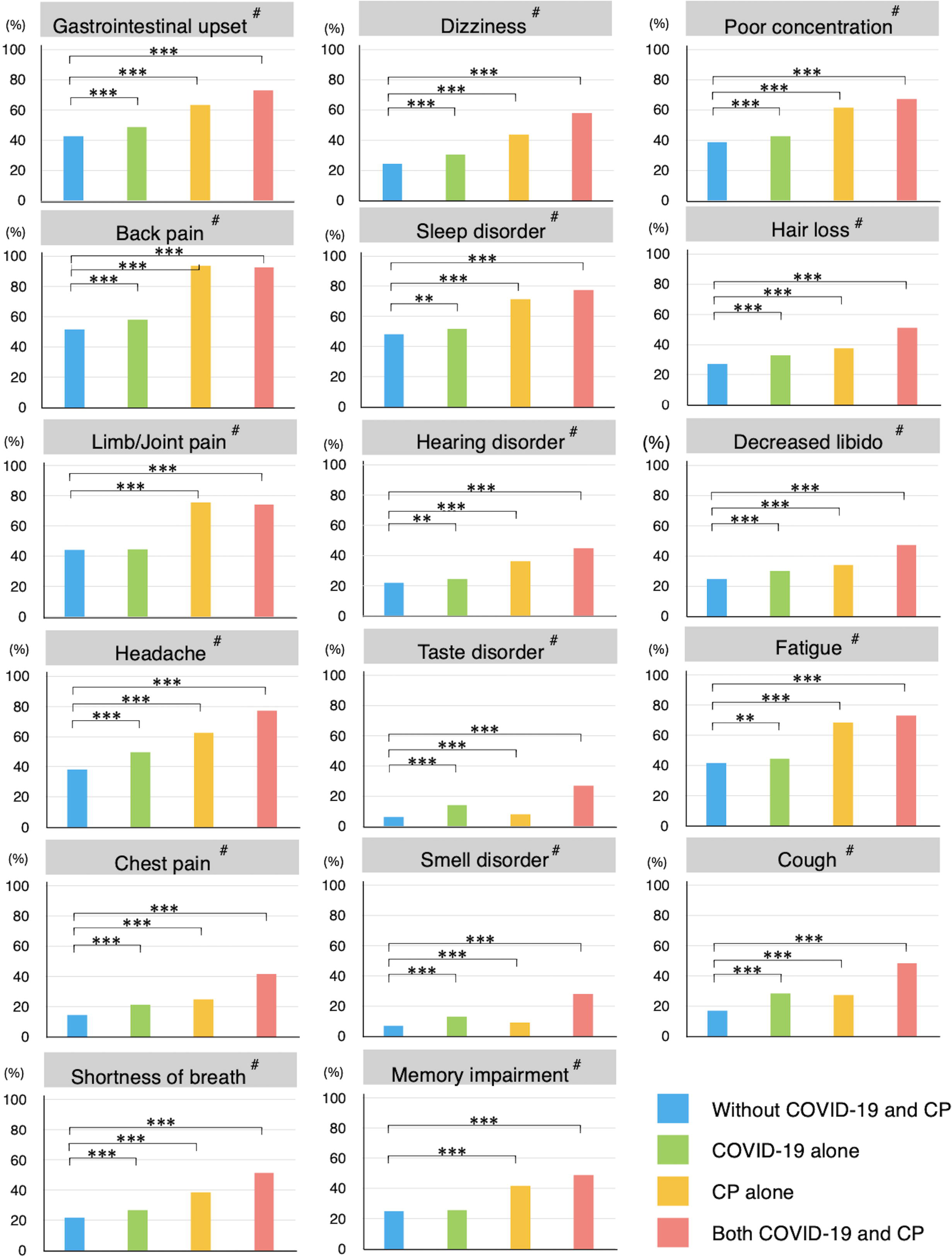
Comparison of the prevalence of each long COVID-like symptom across the groups (N = 28,611). Chi-square tests were performed for the four groups (#p < 0.001). Post hoc comparisons against the group without chronic pain and COVID-19 were performed using the chi-square test (**p < 0.01/3, ***p < 0.001/3.)

Multivariable logistic regression analysis revealed greater prevalence odds for 14 of the 17 symptoms, including gastrointestinal upset, back pain, limb/joint pain, headache, chest pain, shortness of breath, hearing disorder, taste disorder, smell disorder, memory impairment, poor concentration, hair loss, decreased libido, and cough, among individuals with COVID-19 alone than those without CP and COVID-19 (Figure 3). Furthermore, individuals with CP alone exhibited greater prevalence odds for 15 of the 17 symptoms, including gastrointestinal upset, back pain, limb/joint pain, headache, chest pain, shortness of breath, dizziness, sleep disorder, hearing disorder, memory impairment, poor concentration, hair loss, decreased libido, fatigue, and cough, than those without CP and COVID-19. Except for back pain, limb/joint pain, and fatigue, for which people with CP alone showed the highest prevalence odds, those with both CP and COVID-19 exhibited the highest prevalence odds for the other 14 symptoms. Moreover, individuals with both CP and COVID-19 exhibited significantly increased prevalence odds for 9 of the 17 symptoms, including chest pain, shortness of breath, dizziness, hearing disorder, smell disorder, memory impairment, hair loss, decreased libido, and cough, compared to those with either CP or COVID-19 alone.

**Figure 3.**
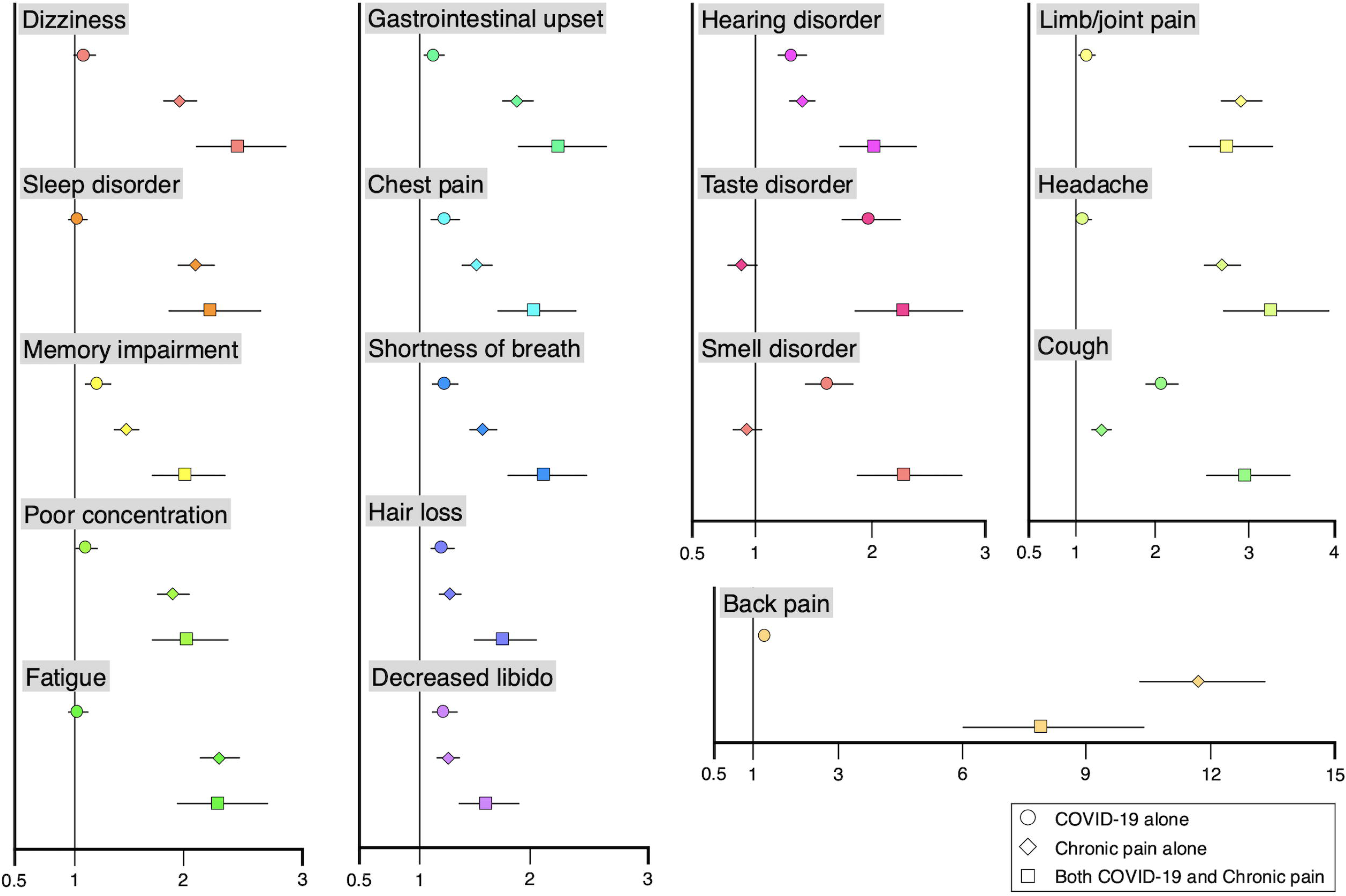
Odds ratios of long COVID-like symptoms for each group (N = 28,611). The odds ratios for the prevalence of each long COVID-like symptom were computed to assess the effect of COVID-19 alone, chronic pain (CP) alone, and both COVID-19 and CP using a multivariable logistic regression analysis. Potential confounding factors associated with either or both chronic pain and long COVID, including age, sex, BMI, employment status, marital status, highest educational level, income, number of comorbidities, psychological distress, number of COVID-19 vaccinations, and smoking habits, were adjusted for the analysis. The bar represents 95% confidence interval.

## Discussion

### Chronic pain and long COVID-like symptoms

Although individuals with CP exhibit various unspecific physical issues, few studies have investigated the actual nature and prevalence of these symptoms. Our study revealed that a total of 5,067 individuals with CP suffer from a greater number of long COVID-like symptoms than those without CP and even those with COVID-19 alone. Furthermore, individuals with CP had a significantly higher prevalence of most of the long COVID-like symptoms investigated in this study.

Previous studies have identified several risk factors for long COVID, including the severity of acute COVID-19 infection, use of invasive mechanical ventilation, older age, female sex, multiple comorbidities, smoking, poor mental health, and lower economic status.^10,16,17^ The present study provides further insights into the epidemiology of long COVID, highlighting that CP is an independent risk factor for various long COVID-like symptoms, which may even surpass the influence of COVID-19 on many of these symptoms. Therefore, the ‘long COVID’ symptoms may not solely represent the long-term effect of COVID-19.

### Association between chronic pain and long COVID: the central sensitisation hypothesis

CP and long COVID exhibit similar symptoms, potentially because of a common underlying mechanism: central sensitisation. Central sensitisation refers to a state in which the central nervous system undergoes neuroplastic changes, leading to the amplification of pain.^19^ It is reported to involve grey matter changes in pain associated brain regions, alteration of brain-network connectivity, and immune system abnormalities.^20,21^ Central sensitisation results in dysfunctions and the excitability of neurons, causing multisensory hyperresponsiveness and subthreshold sensory inputs to elicit pain.^21,22^ This lowering of the global sensory threshold, along with neural dysfunctions, contributes to the development of chronic pain conditions as well as the diverse comorbid symptoms^19,21^ (e.g. fatigue, sleep disorder, mood and cognitive disorder, and sensitivity to external and internal stimuli) that characterise central sensitization syndrome as seen in conditions such as fibromyalgia, chronic pelvic pain, chronic fatigue syndrome, and irritable bowel syndrome.^21,23,24^ Notably, since multisite pain, fatigue, and gastrointestinal disorder are listed as long COVID-like symptoms, central sensitisation may be a common underlying mechanism in both CP and long COVID.

### Current knowledge on mechanisms underlying the long COVID

The pathogenesis of long COVID involves mechanisms such as immune dysregulation, autoimmunity, dysfunctional neurological signalling, microbiota dysbiosis, endothelial abnormalities, blood clotting, and coagulopathy, which can lead to dysautonomia, neuroinflammation, reduced cerebral blood flow, and abnormal gas exchange in the respiratory system.^25^ These alterations in multiple organ systems result in a wide range of symptoms involving the central nervous system, respiratory, gastrointestinal, and reproductive functions.^25^ Although detailed mechanisms underlying long COVID remain insufficiently understood, it is crucial to acknowledge that both CP and long COVID are reported to involve vulnerability of the central nervous and immune systems.

### Interaction between chronic pain and COVID-19

The prevalence odds of multiple long COVID-like symptoms, such as chest pain, shortness of breath, dizziness, hearing disorder, smell disorder, memory impairment, hair loss, decreased libido, and cough, are significantly higher among the individuals with both CP and long COVID. As both CP and long COVID affect the nervous and immune systems, individuals with CP may experience further modifications in their nervous and immune systems triggered by COVID-19 infection.^20,25^ These interactions may lead to heightened sensitivity of the central nervous system and the onset of new long COVID-like symptoms. Therefore, ‘long COVID’ may not simply reflect the direct effects of COVID-19 infection but could also represent the consequences of central nervous system hypersensitivity triggered by this systemic viral infection.

### Limitations

The present study has some limitations that should be acknowledged. First, we investigated the long COVID-like symptoms as symptoms experienced during the previous week. Therefore, the symptoms surveyed do not strictly meet the definition of long COVID,^26^ but we could still evaluate the trends in the presence of these symptoms. Second, we could not distinguish whether CP existed prior to the COVID-19 infection or whether it developed as a symptom of long COVID after the infection. This point needs to be clarified in future longitudinal studies. Third, we did not evaluate CP according to the location. Considering that central sensitisation is generally involved in any CP regardless of the location, the impact of the pain location may not be significant. However, as central sensitisation is typically characterized bywidespread/multisite pain,^21^ number of bodily parts with pain should be considered in the future. Fourth, the history of COVID-19 was self-reported, and the severity of infection was not included in this study. A more accurate understanding of the relationship among COVID-19 infection, long COVID-like symptoms, and CP can be obtained by incorporating objective diagnosis and severity. Finally, not all symptoms of long COVID symptoms could be included, but we included and assessed the principal symptoms in this study.

## Conclusions

This study revealed that 5,067 people with CP exhibited various long COVID-like symptoms. Furthermore, individuals with CP showed a higher prevalence of certain respiratory and central nervous system-related long COVID-like symptoms compared to those with a history of COVID-19 alone. Additionally, individuals with CP experienced a greater overall number of long COVID-like symptoms and had greater prevalence odds for most of the symptoms compared to those without CP or COVID-19. Collectively, these results suggest that CP is an independent risk factor for the long COVID-like symptoms, and that ‘long COVID’ may not be specifically related to COVID-19 alone.

## Author Contributions

ST: Conceived the study, analysed and interpreted the data, drafted the manuscript, and produced the tables and figures. HS: Drafted the manuscript. MK, CT, and YW: Edited the manuscript. TT: Conceived the study and supervised data acquisition. KW: Designed the study, analysed the data, edited the first draft, and supervised the study. KW, SK, TY and TT: Critically revised the manuscript for important intellectual content.

All the authors have read and agreed to the published version of the manuscript.

## Acknowledgement

We thank Dr. Hitoshi Honda (Department of Infectious Diseases, Fujita Health University School of Medicine, Aichi, Japan) for the support. We also thank Editage (www.editage.jp) for English language editing.

## Declaration of Generative AI and AI-assisted Technologies in the Writing Process

We did not use generative AI or AI-assisted technologies during the writing process.

## Data Sharing Statement

The Japan COVID-19 and Society Internet Survey (JACSIS) study dataset is available upon reasonable request to the corresponding author.

